# Moving diagnostics upstream: prehospital blood gas analysis is associated with safe community care and improved patient selection for hospital admission

**DOI:** 10.64898/2026.04.01.26349943

**Authors:** Harald Lux, Johannes Roth, Stefanie Hemmer, Sebastian Lang, Jan-Christoph Lewejohann, Michael Bauer, Jonas Brock, Petra Dickmann

**Affiliations:** Department of Anaesthesiology and Intensive Care Medicine, Jena University Hospital, Jena, Germany; Leibniz Centre for Photonics in Infection Research, Jena, Germany; Thuringian Centre for Quality and Research in Emergency Medicine and Emergency Medical Services (ThuZenQ), Jena, Germany; Department of Emergency Medicine, Jena University Hospital, Jena, Germany; Emergency Medical Services Medical Directorate, City of Jena Fire Brigade, Jena, Germany

## Abstract

**Background:** Emergency departments (EDs) in high-income countries face rising demand, workforce shortages and crowding. We investigated whether prehospital point-of-care blood gas analysis (BGA), used by emergency physicians, is associated with higher ambulatory treatment rates and improved patient selection for hospital admission.

**Methods:** We retrospectively analysed routinely collected data from a pilot implementation of a mobile blood gas analyser in physician-staffed emergency medical services (EMS) in Jena, Germany (July 2023 to May 2024). Adult emergency patients receiving prehospital BGA were compared with propensity score–matched EMS controls without BGA. Primary outcomes were the proportion treated on scene and, among transported patients, the hospital admission rate. Secondary outcomes were 30-day safety among ambulatory patients and associations between BGA parameters and disposition. We used standardised mean differences to assess balance and receiver operating characteristic analysis for lactate thresholds.

**Results:** Of 109 patients receiving prehospital BGA, 98 met inclusion criteria after excluding 9 patients with missing NACA scores, 1 on-scene death and 1 invalid age record; these were matched to 390 controls (total n = 488). Baseline demographics, severity and vital signs were well balanced. Ambulatory treatment was markedly higher in the BGA cohort compared with matched controls (27.6% vs 8.7%; OR 3.98, 95% CI 2.26 to 7.01; p<0.001). No ambulatory BGA patient required ED re-attendance or repeat EMS contact within 30 days. Among transported patients, 58% in the BGA cohort were admitted to hospital, compared with an overall regional ED conversion rate of approximately 30%. Lactate ≥2.6 mmol/L was the most influential parameter for disposition decisions, with elevated lactate and acid–base disturbances strongly associated with transport and admission.

**Conclusion:** Prehospital BGA was associated with fourfold higher ambulatory treatment rates (27.6%) and a twofold higher ED conversion rate among the patients who were transported (58%), indicating improved risk stratification and resource allocation. These findings suggest that integrating objective biochemical data into prehospital assessment may enhance treat-and-refer decision-making and support more efficient use of limited emergency care capacity.

**KEY MESSAGES:** *What is already known on this topic:* Emergency departments face rising demand and workforce shortages, yet prehospital diagnostics beyond basic monitoring remain limited, restricting the evidence base for safe treat-and-refer decisions in EMS.

*What this study adds:* Prehospital blood gas analysis was associated with a fourfold higher rate of ambulatory treatment (27.6% vs 8.7%; OR 3.98) and a twofold higher ED conversion rate among the patients who were transported (58%), with no unplanned ED re-attendances within 30 days among patients managed on scene.

*How this study might affect research, practice or policy:* These findings support evaluation of prehospital BGA as a tool to inform treat-and-refer decisions, though multicentre studies are needed to confirm generalisability and cost-effectiveness before wider implementation.

## INTRODUCTION

EDs in many high-income health systems face sustained crowding driven by rising case volumes, demographic ageing, multimorbidity and workforce shortages, with adverse consequences for clinical outcomes and staff well-being[1–3]. Policy responses increasingly emphasise alternatives to emergency department attendance, including treat-and-refer EMS models and alternatives to default ED transport[4]. Evidence from programmes such as the US Emergency Triage, Treat, and Transport model demonstrates the feasibility of treatment-in-place when supported by appropriate protocols and system design[5].

Safe community-based care requires reliable risk stratification to distinguish patients who can be managed at home from those needing hospital-based diagnostics and monitoring[6]. Point-of-care testing supports this by providing rapid biochemical information that can complement clinical assessment and vital signs. BGA offers immediate insight into acid–base status, oxygenation, lactate and electrolytes, all relevant to respiratory, circulatory and metabolic emergencies[7].

Despite its established role in EDs and intensive care units, BGA is rarely available in EMS, where diagnostics are often limited to history, examination and basic monitoring[7]. Prior prehospital studies have focused mainly on lactate as a mortality predictor rather than on comprehensive BGA panels and their effect on patient pathways or ED utilisation[8–10]. Studies have shown that prehospital BGA can improve diagnostic accuracy[11], and point-of-care testing can reduce clinical decision time[12]. However, the impact of routine prehospital BGA on disposition, safety and ED resource use remains poorly understood.

We hypothesised that the availability of mobile BGA in a physician-staffed EMS system is associated with higher rates of safe ambulatory treatment among lower-risk patients and improved selection of higher-risk patients for hospital admission. Our primary objective was to evaluate whether prehospital BGA is associated with higher rates of treatment on scene and a higher hospital admission rate among transported patients, without compromising short-term safety.

## METHODS

### Study design and setting

We retrospectively analysed data from a pilot implementation of a mobile BGA device in the physician-staffed EMS system of Jena, Germany. A mobile BGA device (epoc Blood Analysis System, Siemens Healthineers) was stationed at the regional dispatch centre and available for emergency physicians to deploy at their discretion, independent of the emergency vehicle used. The evaluation period ran from July 2023 to May 2024. The EMS operates within a mixed urban–rural catchment with standard German physician-led prehospital care structures.

The pilot implementation was conducted as a non-interventional service evaluation of a CE-certified device used within its intended purpose; no study-related interventions were performed on patients. The retrospective data analysis was reviewed and acknowledged by the Ethics Committee of Jena University Hospital (reference 2026-2026-4150-BO-D), which raised no ethical concerns. The study was not classified as a clinical trial. Informed consent was waived owing to the observational nature and use of routinely collected data. Mobile BGA devices were provided by a commercial supplier (Axonlab) without any role in study design, analysis or reporting. This study is reported following STROBE guidelines for observational studies.

### Participants

We included all adult patients (≥18 years) attended by participating EMS teams in whom the emergency physician considered prehospital BGA clinically indicated. Typical indications included acute dyspnoea, haemodynamic instability, suspected sepsis, metabolic derangement and altered mental status. Of 109 patients receiving BGA during the pilot, 98 met inclusion criteria for the primary analysis (exclusions: 9 missing NACA scores, 1 death on scene [NACA 7], 1 invalid age record).

A control group was drawn from EMS cases without BGA using propensity score matching. Matching variables included age, sex, National Advisory Committee for Aeronautics (NACA) severity score and ICD-10 diagnostic category. Nearest-neighbour matching with a 1:4 ratio and a caliper of 0.2 standard deviations of the logit of the propensity score was applied, yielding 390 matched controls and a total matched cohort of 488 patients.

### Procedure and data collection

The procedure consisted of routine use of a mobile BGA analyser at the scene, integrated into standard EMS assessment and management. BGA parameters included pH, partial pressures of carbon dioxide and oxygen, bicarbonate, base excess, sodium, potassium and lactate.

Data were collated from routine EMS documentation and a structured evaluation form. Variables included demographics, presenting complaint, suspected diagnosis, NACA score, vital signs, BGA parameters and disposition. For each BGA case, physicians rated whether the test was helpful for immediate management and identified the most influential parameter for decision-making.

### Outcomes

Primary outcomes were:

1. the proportion of patients treated on scene without hospital transport (ambulatory treatment), and
2. among transported patients, the hospital admission rate (ED conversion rate).

Secondary outcomes were:

- Safety of ambulatory treatment, defined as absence of unplanned ED re-attendance or repeat EMS contact within 30 days.
- Association between BGA parameters and disposition (ambulatory vs transport; admission vs ED discharge).

For ambulatory patients, follow-up at 24 hours, 72 hours and 30 days used EMS and hospital information systems to identify ED re-attendances or repeat EMS contacts. For transported patients, ED records were used to determine hospital admission.

### Statistical analysis

Continuous variables were summarised as medians with IQRs and categorical variables as counts and percentages. We assessed balance between groups using standardised mean differences, with values <0.1 indicating adequate balance. Between-group comparisons used Wilcoxon rank-sum tests for continuous variables and Fisher’s exact or χ² tests for categorical variables as appropriate. Comparisons of BGA parameters between disposition groups (Figure 2B) were exploratory and not adjusted for multiple comparisons.

Receiver operating characteristic analysis was used to evaluate the discriminative performance of lactate for predicting transport decisions, and Youden’s index identified the optimal threshold. Analyses were performed in R (version 4.4.1). To assess robustness, we repeated propensity score matching across a range of caliper widths (0.05 to 0.50) and matching ratios (1:1 to 1:5), and performed multivariable logistic regression adjusting for the same covariates.

### Patient and public involvement

Patients and members of the public were not involved in the design, conduct or reporting of this observational service evaluation. The research question was developed in response to health system and EMS stakeholder concerns about ED crowding and prehospital diagnostics rather than directly from patient input. We plan to disseminate the results to local EMS leadership and patient representatives through service meetings and public-facing summaries.

## RESULTS

### Cohort characteristics

Of 109 patients who received prehospital BGA during the pilot period, 11 were excluded (9 missing NACA scores, 1 on-scene death, 1 invalid age record), leaving 98 for analysis. These patients represented a range of emergency presentations and severities, with NACA scores ranging from 1 to 6. After propensity matching, baseline demographics, NACA severity and vital signs were well balanced between the 98 BGA and 390 control patients (table 1, online supplementary figures 1 and 3).

**Table 1:** Baseline characteristics of the study population.

| Variable | BGA cohort<br>(n=98) | Matched controls<br>(n=390) | p<br>value |
| --- | --- | --- | --- |
| **Demographics** |  |  |  |
| Age, years, median (IQR) | 72.0 (56.2–84.8) | 73.5 (54.0–85.0) | 0.892 |
| Female sex, n (%) | 49 (50.0%) | 185 (47.4%) | 0.733 |
| **Severity** |  |  |  |
| NACA score, n (%) |  |  | 0.869 |
| NACA 1 | 3 (3.1%) | 6 (1.5%) |  |
| NACA 2 | 18 (18.4%) | 67 (17.2%) |  |
| NACA 3 | 33 (33.7%) | 147 (37.7%) |  |
| NACA 4 | 27 (27.6%) | 107 (27.4%) |  |
| NACA 5 | 15 (15.3%) | 53 (13.6%) |  |
| NACA 6 | 2 (2.0%) | 10 (2.6%) |  |
| **ICD-10 diagnostic category, n (%)** |  |  | 0.999 |
| Cardiovascular | 28 (28.6%) | 104 (26.7%) |  |
| Respiratory | 7 (7.1%) | 28 (7.2%) |  |
| Neurological | 7 (7.1%) | 26 (6.7%) |  |
| Metabolic | 6 (6.1%) | 27 (6.9%) |  |
| Infections | 10 (10.2%) | 41 (10.5%) |  |
| Psychiatric | 11 (11.2%) | 39 (10.0%) |  |
| Trauma | 7 (7.1%) | 28 (7.2%) |  |
| Other | 22 (22.4%) | 97 (24.9%) |  |
| **Vital signs on arrival** |  |  |  |
| Heart rate, /min, median (IQR) | 89.5 (73.0–107.0) | 88.5 (74.0–105.0) | 0.947 |
| Systolic BP, mmHg, median (IQR) | 140.0 (128.0–162.0) | 141.0 (119.0–160.0) | 0.427 |
| Diastolic BP, mmHg, median (IQR) | 85.0 (68.0–97.0) | 80.0 (66.2–94.0) | 0.263 |
| SpO<sub>2</sub>, %, median (IQR) | 96.0 (90.8–98.0) | 96.0 (93.0–98.0) | 0.382 |
| Respiratory rate, /min, median (IQR) | 18.0 (14.0–24.0) | 18.0 (15.0–22.0) | 0.914 |
| **Disposition and operational times** |  |  |  |
| Scene time, minutes, median (IQR) | 37.0 (27.5–46.0) | 28.5 (20.8–37.8) | <0.001 |
| Ambulatory treatment on scene, n (%) | 27 (27.6%) | 34 (8.7%) | <0.001 |

BGA=blood gas analysis. BP=blood pressure. IQR=interquartile range. NACA=National Advisory Committee for Aeronautics score. SpO<sub>2</sub>=peripheral oxygen saturation.

Median age was approximately 72 years in the BGA cohort and 73.5 years in controls, with roughly half of each cohort being female. Distributions of NACA categories and initial vital signs (heart rate, blood pressure, oxygen saturation and respiratory rate) were similar, indicating comparable case mix between groups.

**Figure 1:**
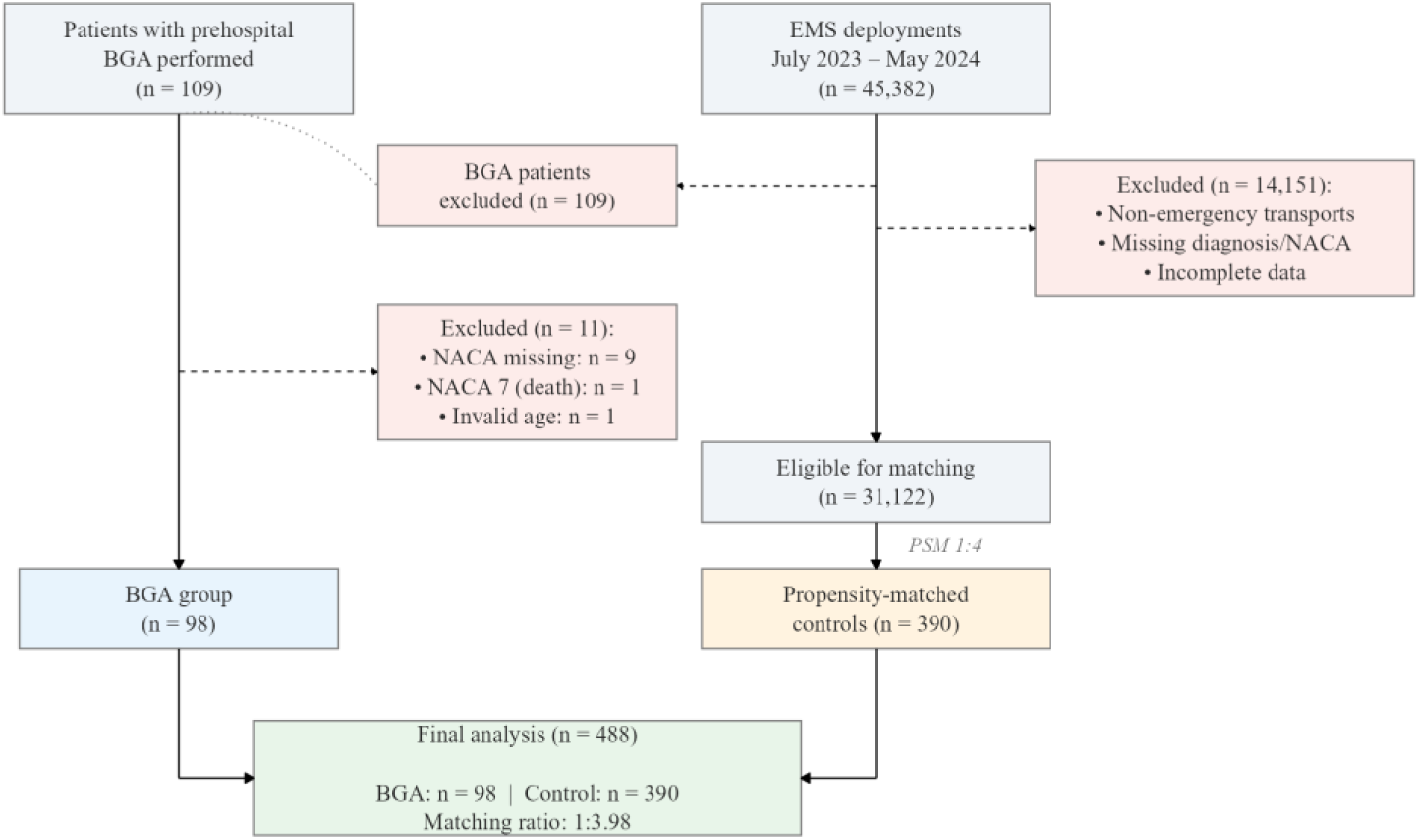
STROBE flow diagram of participant inclusion, exclusions and matching.

### Primary outcomes

Ambulatory treatment on scene occurred in 27.6% of patients in the BGA cohort compared with 8.7% in matched controls (table 2). This corresponded to an OR for ambulatory management of 3.98 (95% CI 2.26 to 7.01; p<0.001).

**Table 2:** Study outcomes.

| Outcome | BGA cohort | Matched controls | OR (95% CI) | p value |
| --- | --- | --- | --- | --- |
| Primary outcome | (n=98) | (n=390) |  |  |
| Ambulatory treatment on scene, n (%) | 27 (27.6%) | 34 (8.7%) | 3.98 (2.26–7.01) | <0.001 |
| **Safety outcomes (ambulatory patients only)** |  |  |  |  |
| Repeat EMS contact within 24h, n (%) | 0 (0.0%) | 2 (5.9%) | — | 0.498 |
| Repeat EMS contact within 72h, n (%) | 0 (0.0%) | 2 (5.9%) | — | 0.498 |
| Repeat EMS contact within 30 days, n (%) | 0 (0.0%) | 3 (8.8%) | — | 0.248 |

BGA=blood gas analysis. CI=confidence interval. EMS=emergency medical services. OR=odds ratio.

Among the 71 transported patients in the BGA cohort, 41 (58%) were admitted to hospital, compared with an overall regional ED conversion rate of approximately 30%[2]. Median scene time was longer in the BGA cohort than in controls (37.0 vs 28.5 minutes; p<0.001; table 1), consistent with the additional time required for point-of-care testing.

### Secondary outcomes

No patient managed ambulatory in the BGA cohort had an unplanned ED re-attendance or repeat EMS contact within 30 days. Increased ambulatory management did not appear to compromise short-term safety in this setting.

Analysis of BGA parameters showed that lactate and acid–base disturbances were strongly associated with disposition (figure 2). Physician feedback forms were completed for 79 of 98 cases (80.6%). Lactate was the single parameter most frequently rated as influential by physicians. A lactate threshold of 2.6 mmol/L was observed to differentiate patients managed in the community from those transported to hospital (AUC 0.66, 95% CI 0.55 to 0.77; sensitivity 85%, specificity 47%). Elevated lactate and marked acid–base derangements were more frequent among transported and admitted patients, supporting their role in risk stratification[9,10,13].

**Figure 2:**
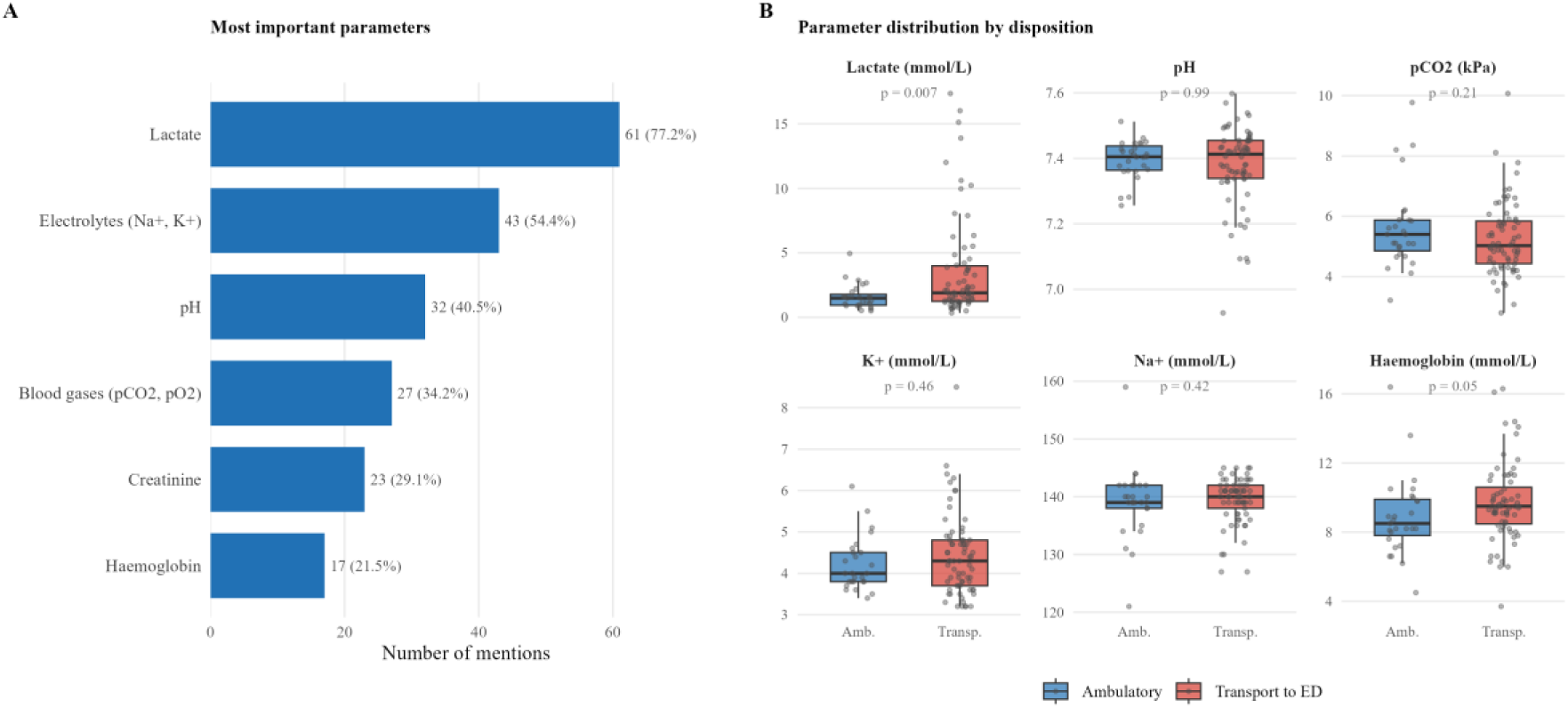
Physician-rated importance of BGA parameters and their distribution by patient disposition. (A) Most important blood gas analysis (BGA) parameters as rated by emergency physicians in the prehospital setting (n=79 responses). (B) Distribution of key BGA parameters stratified by patient disposition: ambulatory treatment on scene versus transport to the emergency department. P values were calculated using the Wilcoxon rank-sum test.

Exploratory subgroup analyses showed that the association between BGA availability and ambulatory treatment was most pronounced among NACA 2 patients (OR 27.0, 95% CI 4.7 to 155.0) and in metabolic presentations (OR 7.4, 95% CI 1.2 to 44.8), with a consistent direction of effect across most diagnostic categories, though subgroup analyses were not pre-specified and individual subgroups were underpowered. These results should be considered hypothesis-generating (online supplementary figure 2).

Sensitivity analyses confirmed the stability of the primary finding (online supplementary table 1). The odds ratio for ambulatory treatment remained stable across caliper widths from 0.05 to 0.50 (OR range 3.76 to 3.93) and across matching ratios from 1:1 to 1:5 (OR range 2.49 to 3.86), with all configurations yielding statistically significant results. Multivariable logistic regression on the full unmatched cohort, adjusting for age, sex, NACA score and diagnostic category, supported the association (adjusted OR 6.14, 95% CI 3.52 to 10.43; p<0.001).

## DISCUSSION

### Principal findings

In this physician-staffed EMS system, prehospital BGA was associated with higher ambulatory treatment on scene and a higher hospital admission rate among transported patients, without evidence of excess short-term adverse events in those managed at home. Objective biochemical data may therefore support both treat-and-refer decisions and risk-based selection for hospital care.

### Comparison with previous work

Prior work on prehospital lactate and point-of-care testing has largely focused on mortality prediction and diagnostic timeliness rather than on patient pathways and ED utilisation[7,9,14].

Our results extend this evidence by linking BGA-supported decision-making to concrete changes in disposition, with more patients safely treated in the community and a higher proportion of transported patients ultimately admitted. This pattern indicates improved risk separation, consistent with policy goals of reducing low-risk ED attendance while protecting access for higher-risk patients[2–4].

The observed 58% conversion rate among transported patients compares favourably with prior reports of lower conversion rates and substantial proportions of non-urgent ED visits[2,3]. This suggests that BGA-supported disposition decisions concentrated transport on patients who were more likely to require hospital-level care. By providing objective biochemical data in addition to vital signs and clinical assessment, prehospital BGA appears to support more confident non-transport decisions and targeted transport of patients with biochemical markers of higher risk[15,16].

### Implications for practice and policy

For clinicians, prehospital BGA provides immediate biochemical data that can strengthen clinical judgement about whether patients can be safely managed at home or require hospital-based care[17]. This may increase confidence in treat-and-refer decisions and reduce defensive transport in borderline cases[18]. For EMS organisations and EDs, wider adoption of prehospital diagnostics could become a means to reduce ED crowding and better align demand with limited capacity[1,4]. From a policy perspective, these findings support efforts to rebalance care towards home and community settings when safe, while maintaining appropriate safeguards for higher-risk patients[4–6]. Prehospital BGA could be integrated into broader emergency care redesign initiatives, including pathways for chronic disease exacerbations, sepsis and respiratory compromise, and should be considered alongside teleconsultation, alternative destinations and new payment models[19].

Future work is needed to determine cost-effectiveness and to define indication criteria for specific presentations. Evaluating scalability across different EMS configurations, including paramedic-led systems, will be particularly important[7,20]. Qualitative work involving patients and EMS clinicians is essential for understanding acceptability and implementation challenges[18].

### Strengths and limitations

Strengths of this evaluation include systematic data collection during routine care, use of propensity score matching to reduce confounding, and combination of process outcomes with 30-day safety follow-up. The study also aligns with current policy priorities on ED decongestion and earlier intervention in the care pathway.

Limitations include the single-centre setting and modest sample size, which may limit generalisability and statistical power for subgroup analyses. BGA indication was at the discretion of emergency physicians, introducing potential selection bias, and the non-randomised, observational design precludes firm causal inference. We did not conduct formal cost-effectiveness analysis or examine patient-reported outcomes. Finally, the physician-staffed EMS model may not reflect the impact of BGA in paramedic-only systems. This study was not prospectively registered in a trial registry, as it was designed as a non-interventional service evaluation rather than a clinical trial.

This study provides evidence that prehospital BGA is associated with higher ambulatory treatment rates and improved risk selection among transported patients. The fourfold difference in ambulatory care, combined with near-doubling of ED conversion rates and absence of observed adverse outcomes among ambulatory BGA patients, suggests that objective biochemical data may inform prehospital disposition decisions.

We are cautious in our interpretation. With 27 ambulatory BGA patients and an observational design, our findings generate hypotheses rather than establish definitive treatment recommendations. It remains unclear whether the observed associations reflect BGA directly influencing clinical decisions, or whether physicians who chose to use BGA were already more inclined towards ambulatory management.

What we can say is that, in this cohort, no ambulatory BGA patient came to harm within 30 days. For a field that has traditionally erred on the side of transport, this finding —however preliminary—warrants further investigation. Multicentre trials with larger samples, cost-effectiveness analyses, and technological refinement are needed to establish the place of prehospital BGA in routine EMS practice.

## Data Availability

Anonymised aggregate data supporting the findings of this study are available upon reasonable request from the corresponding author, subject to approval by the institutional data protection officer, as individual-level emergency medical services data are subject to German data protection regulations

## CONTRIBUTORS

All authors contributed to study conception and design. JR conceived the service evaluation, designed data collection instruments, coordinated field implementation and trained EMS personnel. JR, SH, and SL conducted data collection. HL and JB conducted the statistical analysis, including propensity score matching and sensitivity analyses, and prepared all tables and figures. HL, JB, and PD drafted the manuscript. All authors contributed to critical revision and approved the final version. PD supervised the study. HL and JR contributed equally as joint first authors. JB and PD contributed equally as joint senior authors.

## ACKNOWLEDGMENTS

We thank the emergency physicians and paramedics of the Jena EMS system for their participation in data collection. We acknowledge Axonlab for providing mobile blood gas analysis devices for evaluation purposes.

