## Supplementary Material for "Moving diagnostics upstream: prehospital blood gas analysis is associated with safe community care and improved patient selection for hospital admission"

**Supplementary Figure 1:** Covariate balance before and after propensity score matching (love plot). Dashed lines indicate the conventional threshold of 0.1 for standardised mean differences. All covariates achieved adequate balance after matching.

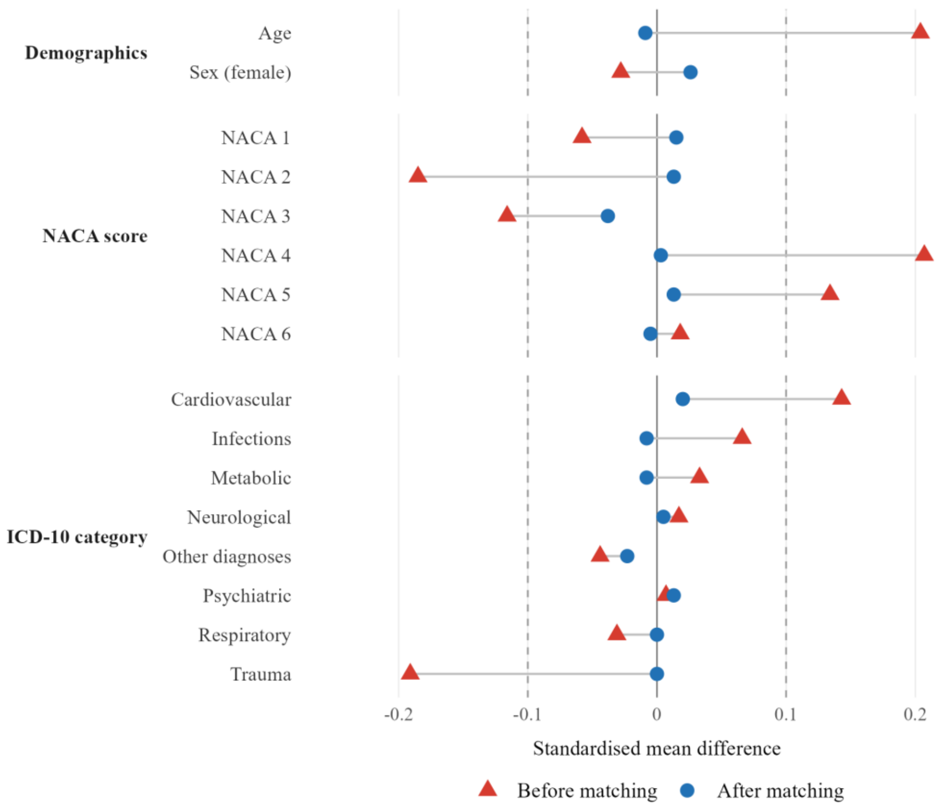

**Supplementary Figure 2:** Forest plot of subgroup analyses for ambulatory treatment by NACA severity score and ICD-10 diagnostic category. Point size is proportional to subgroup sample size. Red points indicate subgroups where the 95% confidence interval excludes 1. All subgroup analyses were exploratory and not pre-specified.

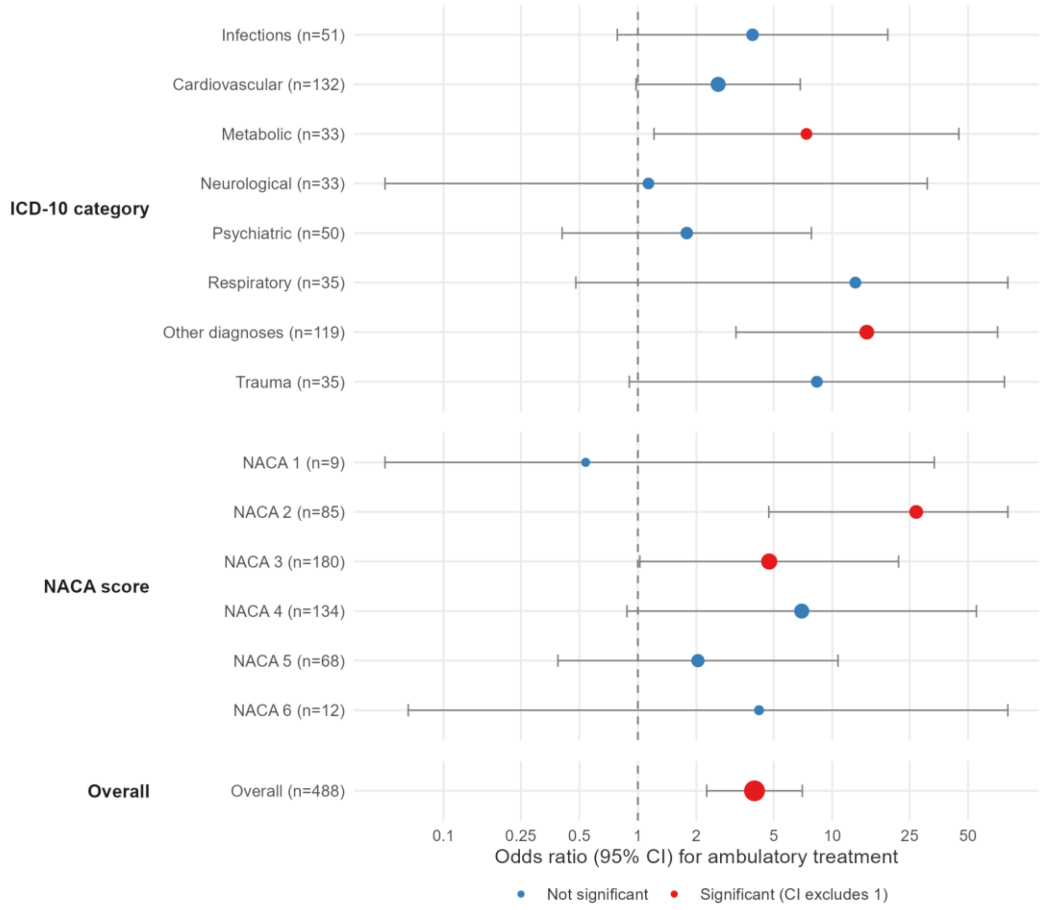

**Supplementary Figure 3:** Propensity score distribution of BGA and control patients after matching. Substantial overlap between groups indicates adequate common support for propensity score matching.

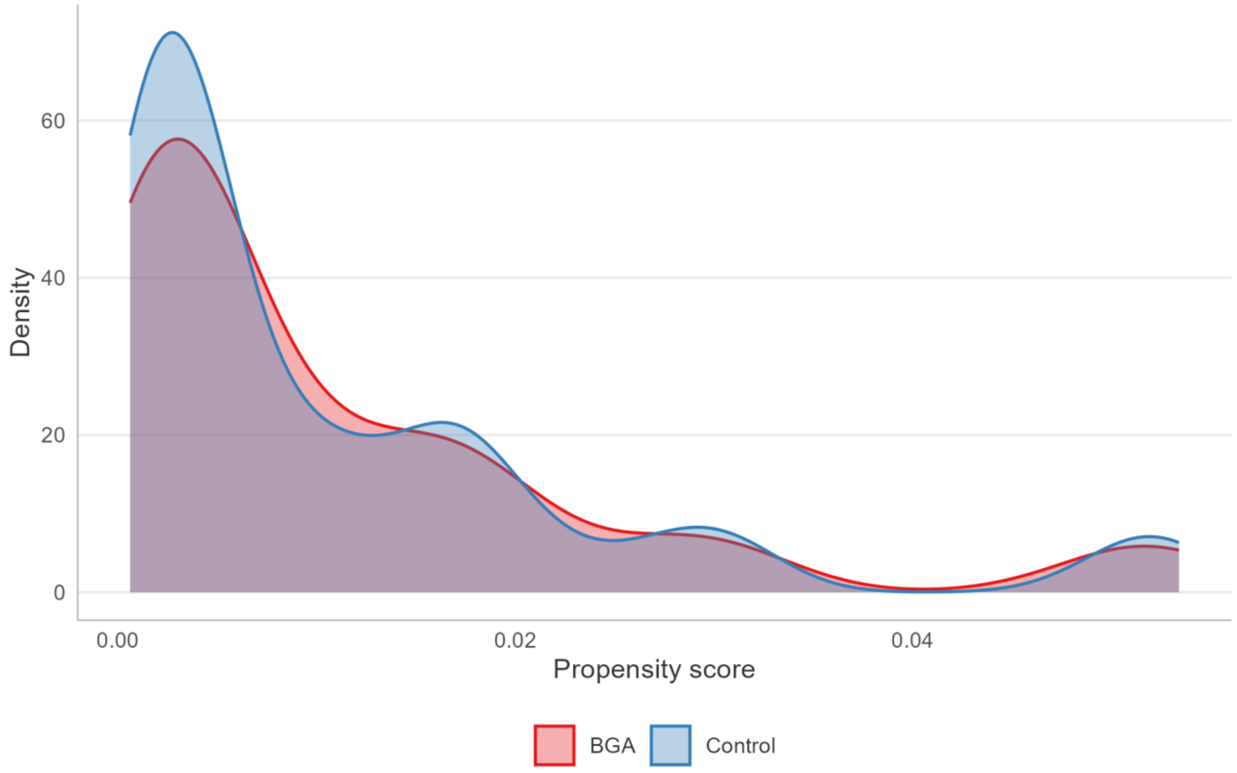

**Supplementary Table 1:** Sensitivity analyses for the association between prehospital BGA and ambulatory treatment.

| Analysis | Specification | OR | 95% CI | p value | n BGA | n Control |
|---|---|---|---|---|---|---|
| **Primary analysis (reference)** | **PSM, caliper 0.2, ratio 1:4** | **3.98** | **2.26–7.01** | **1.12e-06** | **98** | **390** |
| Caliper variation | Caliper = 0.05 | 3.93 | 2.23–6.91 | 1.51e-06 | 98 | 385 |
| Caliper variation | Caliper = 0.1 | 3.85 | 2.19–6.75 | 1.98e-06 | 98 | 389 |
| Caliper variation | Caliper = 0.15 | 3.85 | 2.19–6.75 | 1.98e-06 | 98 | 389 |
| Caliper variation | Caliper = 0.2 | 3.86 | 2.2–6.77 | 1.87e-06 | 98 | 390 |
| Caliper variation | Caliper = 0.25 | 3.86 | 2.2–6.77 | 1.87e-06 | 98 | 390 |
| Caliper variation | Caliper = 0.3 | 3.76 | 2.15–6.59 | 2.73e-06 | 98 | 392 |
| Caliper variation | Caliper = 0.5 | 3.76 | 2.15–6.59 | 2.73e-06 | 98 | 392 |
| Matching ratio variation | 1:1 | 2.49 | 1.19–5.17 | 2.12e-02 | 98 | 98 |
| Matching ratio variation | 1:2 | 3.35 | 1.76–6.35 | 2.55e-04 | 98 | 196 |
| Matching ratio variation | 1:3 | 3.6 | 2–6.49 | 1.98e-05 | 98 | 293 |
| Matching ratio variation | 1:4 | 3.86 | 2.2–6.77 | 1.87e-06 | 98 | 390 |
| Matching ratio variation | 1:5 | 3.82 | 2.22–6.56 | 7.72e-07 | 98 | 486 |

| | | | | | | |
|---|---|---|---|---|---|---|
| Multivariable logistic regression | Unmatched cohort, adjusted* | 6.14 | 3.52–10.43 | 5.34e-11 | 98 | 31122 |

* *Adjusted for age, sex, NACA score and ICD-10 diagnostic category.*
